# Identification of Key Biomarkers for Early Warning of Diabetic Retinopathy Using BP Neural Network Algorithm and Hierarchical Clustering Analysis

**DOI:** 10.1101/2023.05.28.23290657

**Authors:** Peiyu Li, Hui Wang, Zhihui Fan, Guo Tian

## Abstract

**Background:** Diabetic retinopathy is one of the most common microangiopathy in diabetes, essentially caused by abnormal blood glucose metabolism resulting from insufficient insulin secretion or reduced insulin activity. Epidemiological survey results show that about one third of diabetes patients have signs of diabetic retinopathy, and another third may suffer from serious retinopathy that threatens vision. However, the pathogenesis of diabetic retinopathy is still unclear, and there is no systematic method to detect the onset of the disease and effectively predict its occurrence.

**Methods:** In this study, we used medical detection data from diabetic retinopathy patients to determine key biomarkers that induce disease onset through BP neural network algorithm and hierarchical clustering analysis, ultimately obtaining early warning signals of the disease.

**Results:** The key markers that induce diabetic retinopathy have been detected, which can also be used to explore the induction mechanism of disease occurrence and deliver strong warning signal before disease occurrence. We found that multiple clinical indicators that form key markers, such as glycated hemoglobin, serum uric acid, alanine aminotransferase are closely related to the occurrence of the disease. They respectively induced disease from the aspects of the individual lipid metabolism, cell oxidation reduction, bone metabolism and bone resorption and cell function of blood coagulation.

**Conclusions:** The key markers that induce diabetic retinopathy complications do not act independently, but form a complete module to coordinate and work together before the onset of the disease, and transmit a strong warning signal. The key markers detected by this algorithm are more sensitive and effective in the early warning of disease. Hence, a new method related to key markers is proposed for the study of diabetic microvascular lesions. In clinical prediction and diagnosis, doctors can use key markers to give early warning of individual diseases and make early intervention.

## 1. Introduction

According to the ninth edition of Diabetes Overview released by the International Diabetes Federation, 463 million adults worldwide are living with Diabetes, and the total number is expected to rise to 578 million by 2030, with the current global Diabetes prevalence reaching 9.3 percent. Diabetic retinopathy is the most common ocular complication of diabetes, usually occurring 5 years after the onset of type 1 diabetes and 20 years after the onset of type 2 diabetes, which has become the primary reason of blindness worldwide [1-3]. Diabetic retinopathy is a chronic disease where the retina blood vessels of patients are damaged due to high blood sugar, leading to retinopathy. These damages can cause symptoms such as vascular leakage, neovascularization, retinal edema, and bleeding, ultimately resulting in vision impairment and blindness. It is estimated that over 9.3 million people worldwide have become blind due to diabetic retinopathy, with about 90% of cases being preventable. Therefore, predicting and preventing diabetic retinopathy has become a global public health issue that requires enhanced research and practice.

Up to now, diabetes has become the third chronic disease, which is after cardiovascular diseases, malignant tumor diseases. Diabetes is a clinically heterogeneous glucose intolerance syndrome, which is mainly due to the selectivity of immune mediated islet beta cell damage caused by a lack of insulin and glucose metabolism disorders. Chronic hyperglycemia in the body poses serious health risks, and leads to a form of retinopathy, which is known as diabetic retinopathy [4]. Compared with the research on type 1 diabetes and type 2 diabetes, the current studies on the pathogenesis and early warning of diabetes complications are relatively rare. Some scholars have been dedicated to researching the risk factors and related genes that lead to diabetic retinopathy [5-6], and the prediction of diabetic retinopathy through retinal images [7-9]. Yun J et al. investigate the metabolic features of diabetic retinopathy by using metabolomics profiling [10]. The study found significant differences in the concentrations of metabolites among different groups of diabetic retinopathies, and 16 metabolites were identified as common metabolites of non-proliferative diabetic retinopathy (NPDR) and proliferative diabetic retinopathy (PDR), among which three metabolites were found to be potential markers of diabetic retinopathy progression. The identification of these metabolic features will help to deepen our understanding of the mechanisms underlying diabetic retinopathy and provide a basis for the prevention and treatment of related diseases. Carmen et al. construct the linear relationship between the graduated indicators of the diabetes risk assessment model according to the theory of physique identification [11]. They investigated the feasibility of using complexity analysis of glucose profile to predict the development of type 2 diabetes in high-risk individuals. And they found that the complexity metric (detrended fluctuation analysis, DFA) calculated from 24-hour glucose time series using DFA can significantly predict the development of type 2 diabetes in high-risk individuals. The results showed that DFA is a significant predictor of type 2 diabetes development, even after adjusting for other clinical and biochemical variables. This method has the potential to identify patients in need of intensified treatment and provide new insights for the prevention and management of diabetes. Shankar et al. constructed a diabetes risk assessment model to study the linear relationship between biochemical indicators, and proposes a deep learning-based automated detection and classification model for fundus diabetic retinopathy images [12]. Chakravarthy et al. discussed the diagnostic efficiency and accuracy of diabetic retinopathy based on artificial intelligence system, and proposed a framework DR-NET using stacked convolutional neural networks for diabetic retinopathy detection from digital fundus images [13]. Progress in artificial intelligence for diabetic retinopathy screening, including artificial intelligence applications in ‘real-world settings’ are summarized in Gunasekeran’s research. The use of artificial intelligence models for diabetic retinopathy risk stratification, improved the efficiency of diabetic retinopathy management [14]. Somasundaram designed a Machine Learning Bagging Ensemble Classifier to identify retinal features for diabetic retinopathy disease diagnosis and early detection using machine learning and ensemble classification method [15]. However, the previous studies on the risk warning of diabetic complications usually relied on the detection of a large number of biochemical indicators or gene sequences, which requires a more complicated detection and diagnosis process.

In order to reduce the cost of diabetes detection and save medical resources, an increasing number of studies are using artificial intelligence models for early warning of diabetic retinopathy risk, such as convolutional neural networks [16], deep neural networks [17-18], deep learning [19], BP neural networks [20], and so on. In this paper, we constructed a diabetes risk warning model based on the improved BP neural network to determine the key markers affecting the onset of diabetes. Artificial Neural Network (ANN) was first proposed in 1943 and has been widely used in medical diagnosis, prognosis, survival analysis, clinical decision-making and other medical fields. BP neural network is a highly nonlinear mapping network, which can reveal the nonlinear relationship between the medical diagnostic indexes of T1DM patients. In recent years, scholars have applied neural network to the implementation prediction of blood glucose in insulin-dependent diabetes patients, and proved that its prediction effect is superior to other methods [21]. Su B established a prediction model by analyzing the relationship between diabetic retinopathy and related metabolic and biochemical indicators, and the experimental results showed that the model had a high accuracy in diabetes risk assessment [22]. This technology can help people to study and treat diseases by mining hidden and valuable information from the existing medical data. Moreover, it has few applications in medical treatment, especially in diabetes complications, so it is worth exploring its potential value in depth.

Currently, some studies have used convolutional neural network algorithms, data-driven methods, or embedded deep learning to construct diabetes detection models [23-24]. However, there is little research on early warning of diabetic retinopathy risk. Based on this, we will focus on the study of key markers of diabetic retinopathy and early warning of diabetes risk. Based on BP neural network theory, a new diabetes early warning model was established, which could predict the onset of diabetes by identifying the index value of key markers of the disease. In clinical practice, we only need to detect the key markers and calculate the risk warning value of the key markers by using the algorithm, so as to further improve the identification of the key early warning factors in the complications of diabetes, and provide more references and basis for the scientific diagnosis and treatment of the disease. In the research process, the real patient data comes from PHDA, a data warehouse of the National Population Health Science Data Center of China.

## 2. Materials and Methods

### 2.1 Data

In this paper, the Data is from PHDA, a data warehouse of the National Population Health Science Data Center of China (NPHDC), CSTR: A0006.11. A0005.202006.001018. The study object in this paper was 200 patients with diabetic retinopathy, among which the first 180 patients were sampled as the training set, and the last 20 patients were set as the test set. Statistics shows there were 125 male cases and 75 female cases. The data set contained 70 test indicators, By eliminating loss serious, strong noise in data index, the final data set involves 68 items disease research indexes, including 41 items basic physique indexes in patients and 27 items laboratory testing indexes (19 items blood biochemical, 4 items of routine blood, 4 items blood coagulation. Due to lack of data set for a small amount of sample, we use K - Nearest Neighbor (KNN) algorithm to interpolate the missing data, and do minimum - maximum standardized processing for the integrity sample data. The data set contains 200 samples and each sample consists of 28 input parameters and 1 output parameter. In the following, the calculation will be done based on the standardized sample data.

### 2.2 Improved BP neural network theory

BP neural network can be used to study the relationship between the detection indexes and the risk of patients with diabetes complications. When the sample size or research indexes are large, the model fitting effect can be best achieved by adjusting the internal parameters of the algorithm, and the warning effect can be more reliable. The improved architecture of the BP neural network is shown in Figure 1.

**Figure 1.**
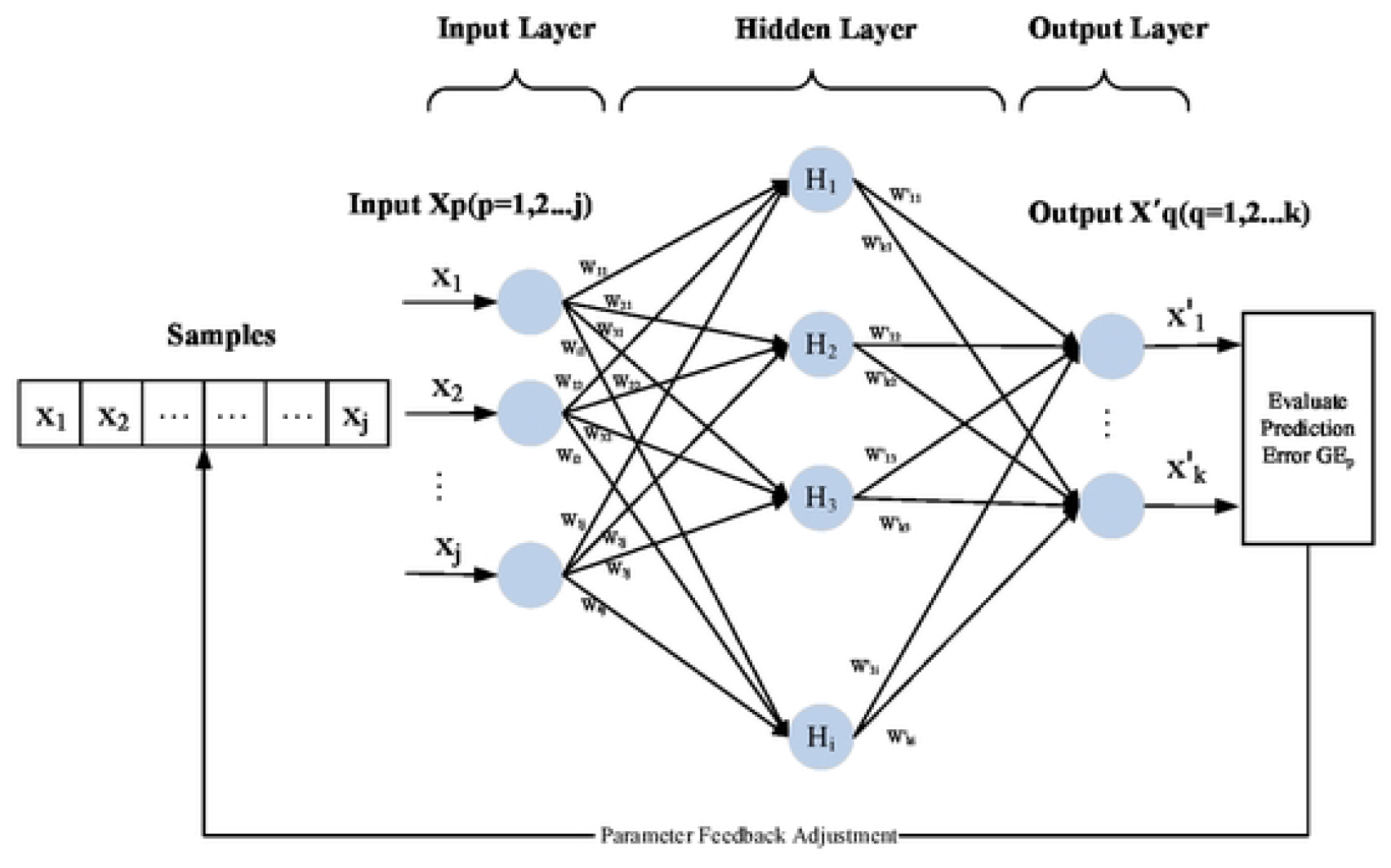
Improved BP neural network Architecture

The 28 indicators data of diabetic patients were taken as input samples, and fasting glucose (GLU) was taken as output value. Specifically, the network includes 3 layers, namely, the input layer, the hidden layer and the output layer. The input parameter was set as *X*_*p*_(*p* = 1,2,⋯,*j*), and the output parameter was 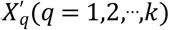. The linear transformation of *X*_*p*_ was carried out to obtain the input *HI*_*i*_ and output *HO*_*i*_ of each node of the hidden layer, which changed as follows

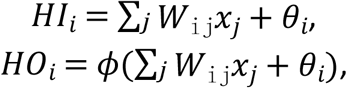

where *W*_ij_ is the connection weight of the *i*th node of the hidden layer reaching the *j*th node of the input layer, *ϕ* is the excitation function of neural network. We choose the Sigmoid function, i.e., 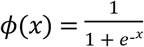. In which *x*_*j*_ represents the training data of the *j*th node in the input layer, *θ*_*i*_ is the node threshold of the hidden layer.

When building a network, we try to use different node transfer function, training function, network learning function and performance analysis function. By comparing the network prediction errors of different settings, we can find when the prediction accuracy is highest, the function setting is as follows, The s-type functions ‘tansig’ and’’lodsig’ are taken as node transfer functions, the momentum inverse gradient descent algorithm ‘TrainLM’ is taken as training function, the function driving quantity term ‘trainlm’ is taken as learning function, and the mean square error MSE is taken as network performance analysis function.

After the training and feedback adjustment of layers of nodes, the result *O*_*k*_ of output nodes is obtained as follows

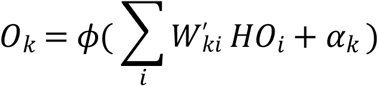

where 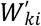 is the connection weight between the *k*th node of the output layer and the *i*th node of the hidden layer, and *α*_*k*_ is the node threshold of the output layer. *D*_*k*_(*x*_*p*_) is the actual value of the predicted sample node at the output layer, and the prediction error *GE*_*p*_ is used to test the prediction accuracy of the model.

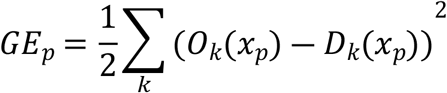

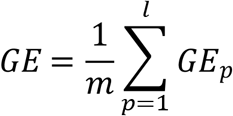

### 2.3 Disease early warning based on key markers

Through training for data by BP neural network, when network prediction rate reaches the optimal level, we record the weight of each index when signals transfer from input layer to the hidden layer. Next, we conduct hierarchical cluster analysis on the weights of these indexes and extract the higher-weight indexes from all indexes. Then, we study the internal correlation of higher-weight indexes and their correlation with low-weight indexes. Spearman correlation coefficient is conducted according to the following formula

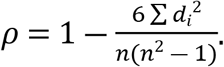

These higher-weight indexes are the key markers of disease warning that we studied. The traditional way to test for diabetes is to detect fasting venous blood glucose and 2H venous blood glucose (OGTT) by a single method, and key markers can be used as a new method to systematically warn the occurrence of such complex diseases.

In order to obtain strong warning signals for disease surveillance, we constructed the following warning index EWI,

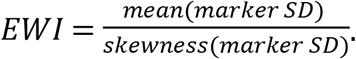

where marker SD refers to the score matrix of the key markers after standardization, which is represented by y_ij_, 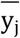 is the column average of the standardized scores of the key markers; Skewness marker SD represents the sample inclination of the standardized score of the key marker and is represented by s_j_.

(1) **Standardized processing:** The original data of key markers are standardized according to rows to eliminate the dimensional influence between different index data, which is specifically calculated as follows

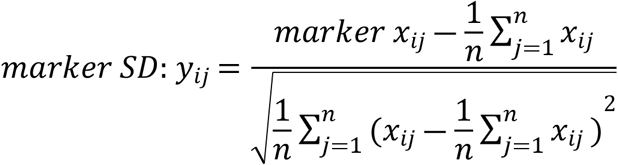

(2) **Arithmetic averaging:** the standardized score matrix is processed by column averaging to obtain 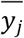, i.e., the comprehensive index of different individuals is calculated as follows

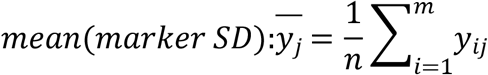

(3) **De-skew processing:** The data skew of the key marker index data of a single individual is calculated, which is a key step to enhance the sensitivity of the warning index to the data fluctuation. It is calculated as follows

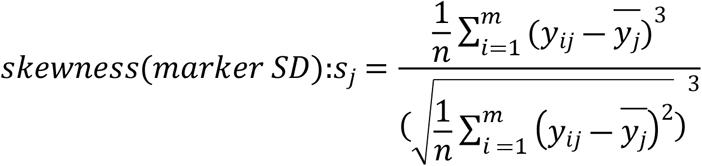

As a key marker, EWI can show drastic fluctuations when individual indexes are abnormal, which is called strong warning signal. When a strong warning signal occurs, the correlation value between key markers is generally high, and much higher than the correlation coefficient between internal indexes of key markers and other indexes. This feature can be used as an important feature in identifying key markers. At the same time, when key markers are used for disease warning, EWI index can show stronger fluctuations than traditional detection indexes, and it will give a strong warning signal when there are abnormalities in individual sign data.

## 3. Results

### 3.1 Model simulation based on BP neural network

The BP neural network model given in Section 2 has been used to train the data mentioned above (https://www.ncmi.cn/). See the appendix for detailed code.

By using BP neural network, we can research risk early warning of diabetic retinopathy (CSTR: A0006.11 A0005.202006.001018) from 28 indexes, such as glycosylated hemoglobin, hemoglobin and triglyceride, total cholesterol, fasting glucose, and so on. These indexes come from the medical data of patients with diabetic retinopathy. The data is divided into training set and test set at a ratio of 75% to 25%, and the output of the network is GLU index value.

The model can predict the change trend of GLU in patients based on the data of 28 medical indexes detected clinically, and train the network with 75% of the data. When the model meets the preset accuracy, the remaining 25% of the data is used to test the performance of the model, as shown in Figure2.

**Figure 2.**
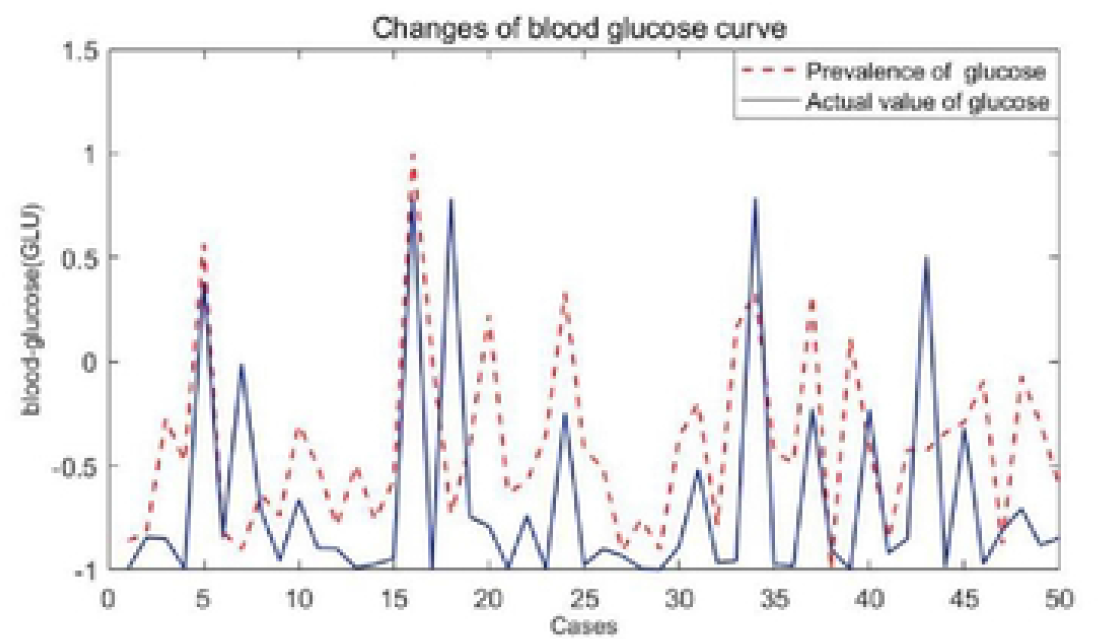
Comparison of predicted value and actual value based on BP neural network

Figure2 shows the predicted and actual values of fasting glucose in 200 medical samples. We can see that the blue line and the red line in the figure almost coincide, indicating that the model can well predict the GLU level of individuals. Figure3 shows the prediction error level of the model, with its relative error rate floating around the 0 level. The two figures show that the data fitting effect of this method is very good, and the prediction error is relatively low.

**Figure 3.**
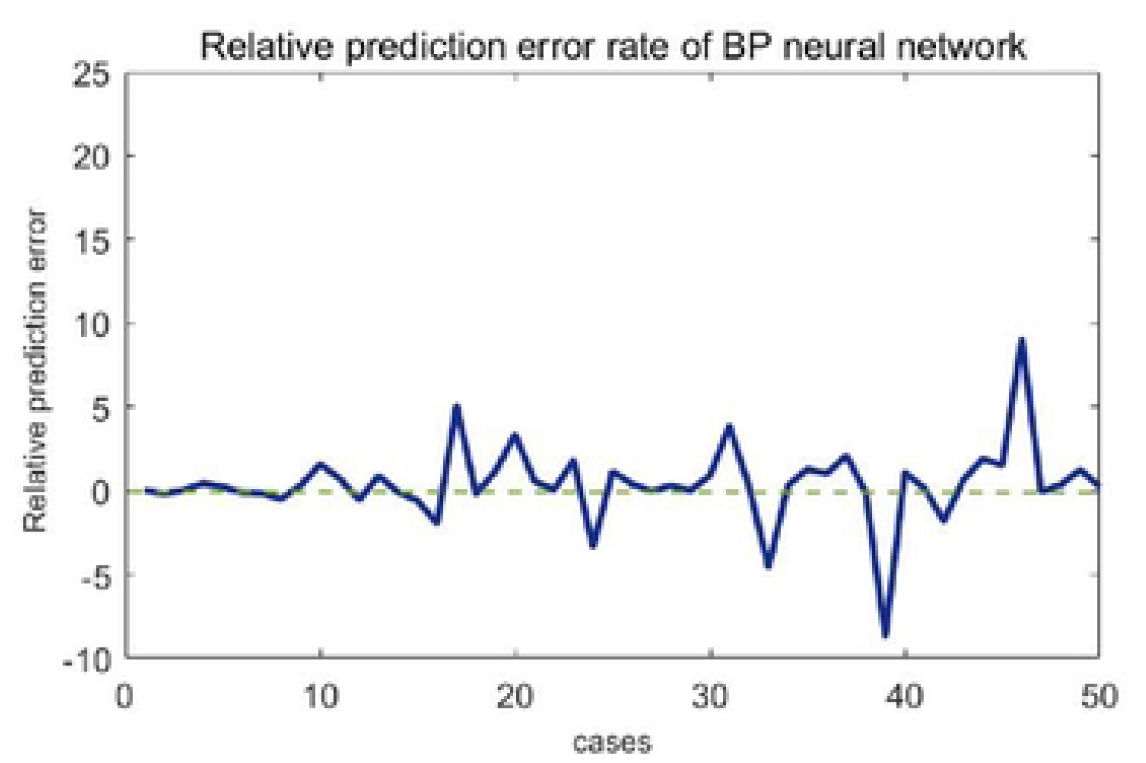
Relative prediction error rate of fasting glucose based on neural network

In order to further detect the key markers affecting the occurrence of diabetic complications (retinopathy), we carried out an in-depth study on the weight matrix of the neural network transmission layer. First, a three-dimensional curved graph was used to show the weight relationship between 28 biochemical indicators, and the results were shown in Figure 4.

**Figure 4.**
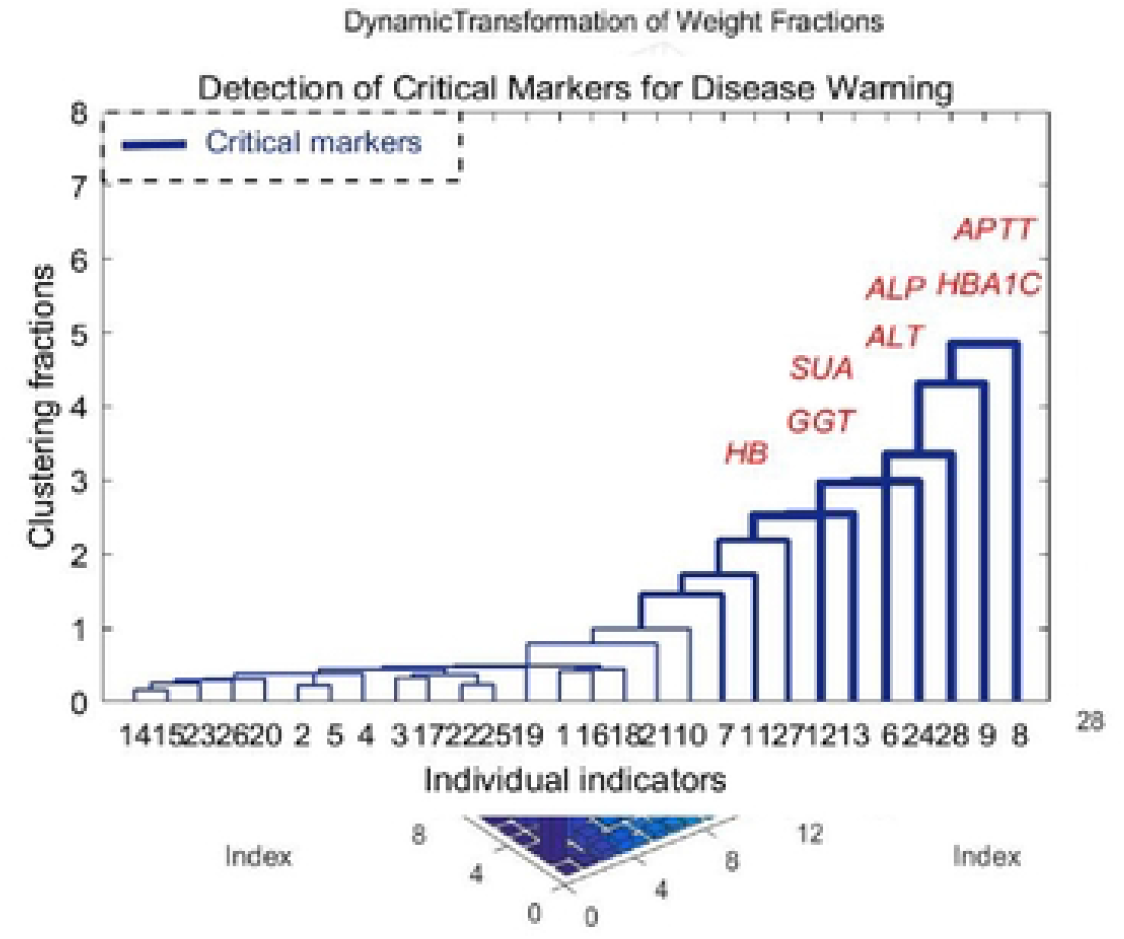
Significance recognition of indicators of diabetic retinopathy

In Figure 4, straight square column presents the importance of the various indicators in the prediction of individual GLU curve. The figure shows that the higher weight of biochemical indexes include glycosylated hemoglobin (HBA1C), total cholesterol (TC), total protein (ALB). Glycosylated hemoglobin is the product of combining hemoglobin in red blood cells with sugars in serum, and is also commonly used as a test indicator for controlling diabetes. The TC value of total cholesterol is an independent risk factor for diabetic retinopathy (DR). The investigation results showed that the blood lipid index level could be timely adjusted according to the TC value of DR patients in clinical practice to prevent the occurrence of DR. Serum total protein can be divided into albumin and erythropoietin and has the physiological function of transporting a variety of metabolites and regulating the transported substances. However, the occurrence of diabetes is not caused by a single factor, but by the joint action of multiple indexes that constitute the key markers. Therefore, we used the weight matrix to further study the key markers for disease early warning.

### 3.2 Detection of key markers for early warning of diabetic complications

Hierarchical clustering was used to analyze the weight data of 28 indicators. The weight data were arranged in descending order and the Euclide distance between different indicators was calculated. The shortest distance method was used to generate the cluster tree. The cluster distance located in the top 5% formed a new module, as shown in Figure 5.

**Figure 5.**
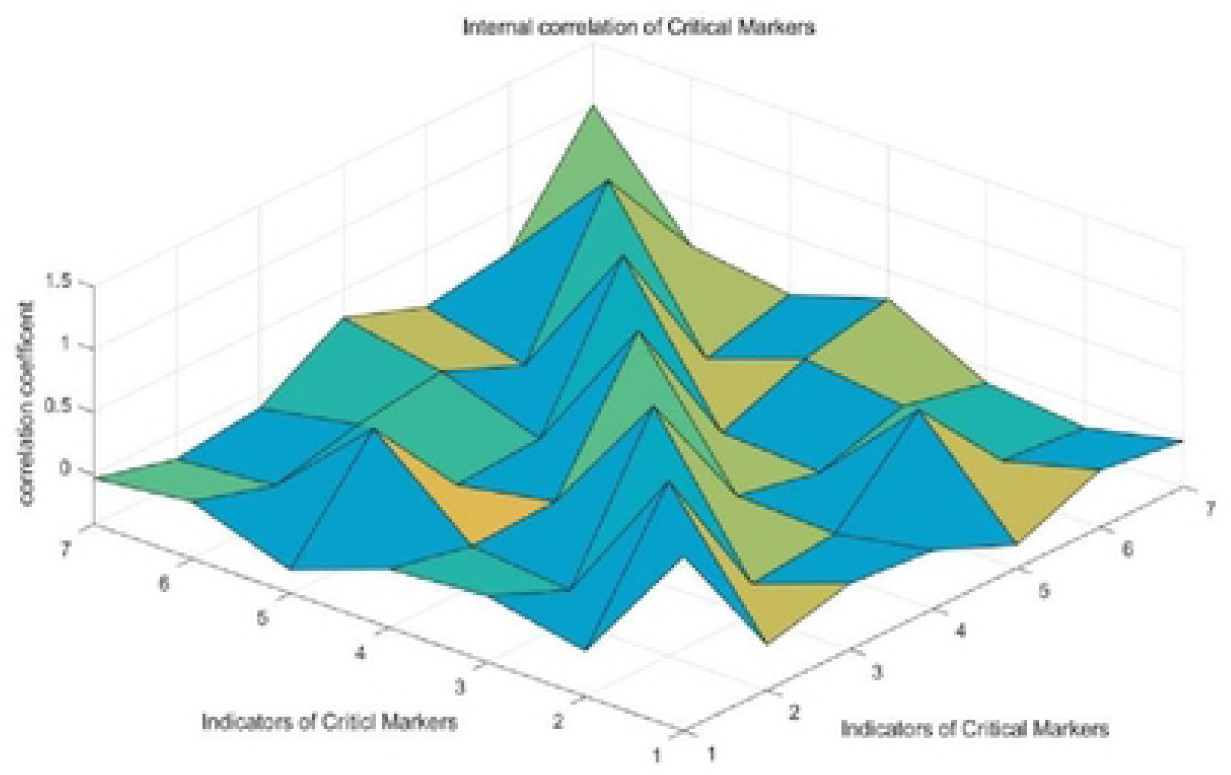
Detection of key markers that induce diabetic retinopathy

Figure 5 shows the hierarchical clustering results, the ordinate is the clustering distance between the weights of biochemical indexes, and the abscess is the classification of biochemical indexes. The new module after clustering consists of 7 factors, namely, glycosylated hemoglobin, serum uric acid, hemoglobin, alanine aminotransferase, glutamine transferase, alkaline phosphatase, and activated partial thromboplastin time (APTT). Changes in HbA1c level can affect the oxygen saturation of motor and venous blood in diabetic retinopathy, and long-term control of blood glucose may delay the progression of DR lesions. Serum uric acid is the final product of the catabolism of purine compounds, which reflects the efficiency of metabolism and decomposition of accounting and other purine compounds to some extent. Hemoglobin is a protein responsible for carrying oxygen in higher organisms. It can form hemoglobin A1c in contact with blood sugar, thus serving as an effective indicator for the detection of diabetes. Its importance has also been successfully verified in our key marker detection. Alanine aminotransferase has also been found to have a good effect in improving lipid metabolism, and has a synergistic effect with AST level. γ-Gamma-glutamine transferase is a kind of oxidoreductase with glutathione function. Its elevated level may be the result of oxidative stress in the body of individuals, which can be used to indicate the loss of oxygen free radical activity to cells. Serum alkaline phosphatase mainly reflects whether the metabolic function of three substances in the body is normal. Studies have shown that bone specific alkaline phosphatase (BAP) is an important indicator of bone metabolism, and anti-tartrate acid phosphatase (BTRACP-5B) is a marker enzyme reflecting osteoclast activity and bone resorption. Therefore, biochemical indicators of bone conversion can be used as a reference for predicting diabetic retinopathy and its severity. PTT is one of the indicators that can reflect an individual’s clotting function. Hyperglycemia can form glycosylation modifications on thrombin, leading to activation of the clotting mechanism. Shortening of APTT is considered a marker of hypercoagulability. Research has shown that the relationship between diabetic retinopathy and the state of coagulation activation can be understood by detecting the changes of coagulation function in diabetic patients. All the above important indicators related to diabetic retinopathy were detected in the key markers. The initiation of diabetic retinopathy is mainly induced by individual lipid metabolism, cell REDOX, influence on bone metabolism, bone resorption and cell coagulation function.

### 3.3 Characteristic analysis of key markers

In order to further verify the early-warning effect of key markers on disease, correlation analysis was conducted for the indicators identified as key markers. The spearman correlation coefficient between internal indicators of key markers and non-marker indicators was calculated respectively. The results were shown in Figure 6 and Figure7

**Figure 6.**
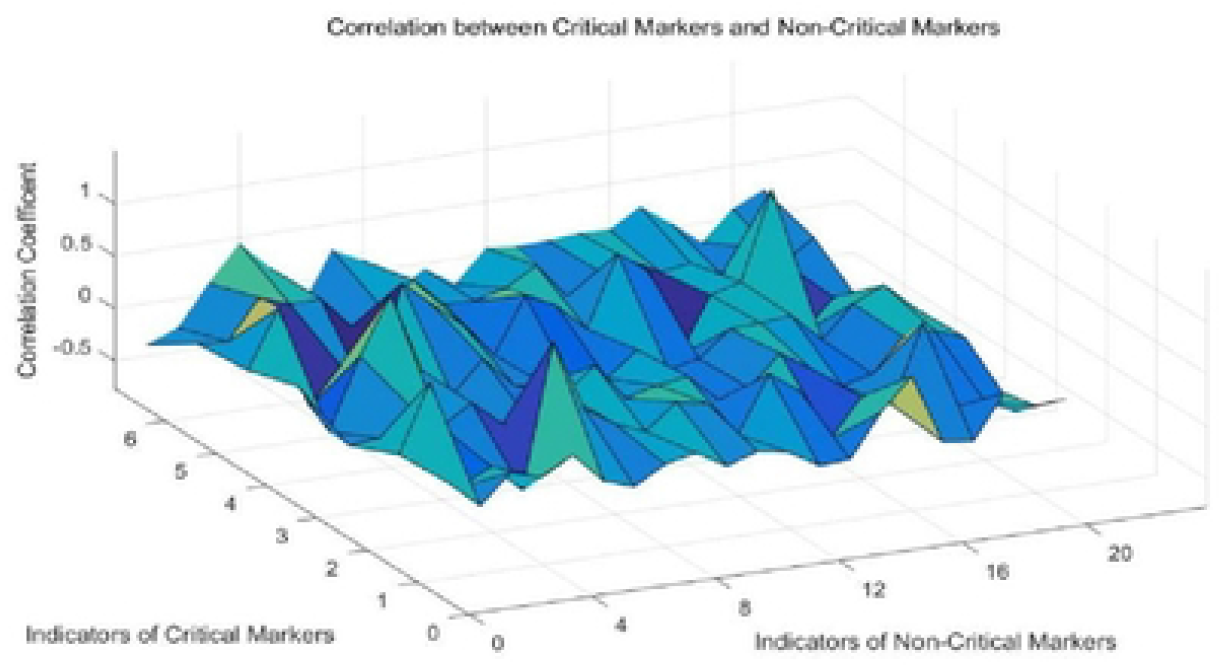
Internal correlation of key markers

**Figure 7.**
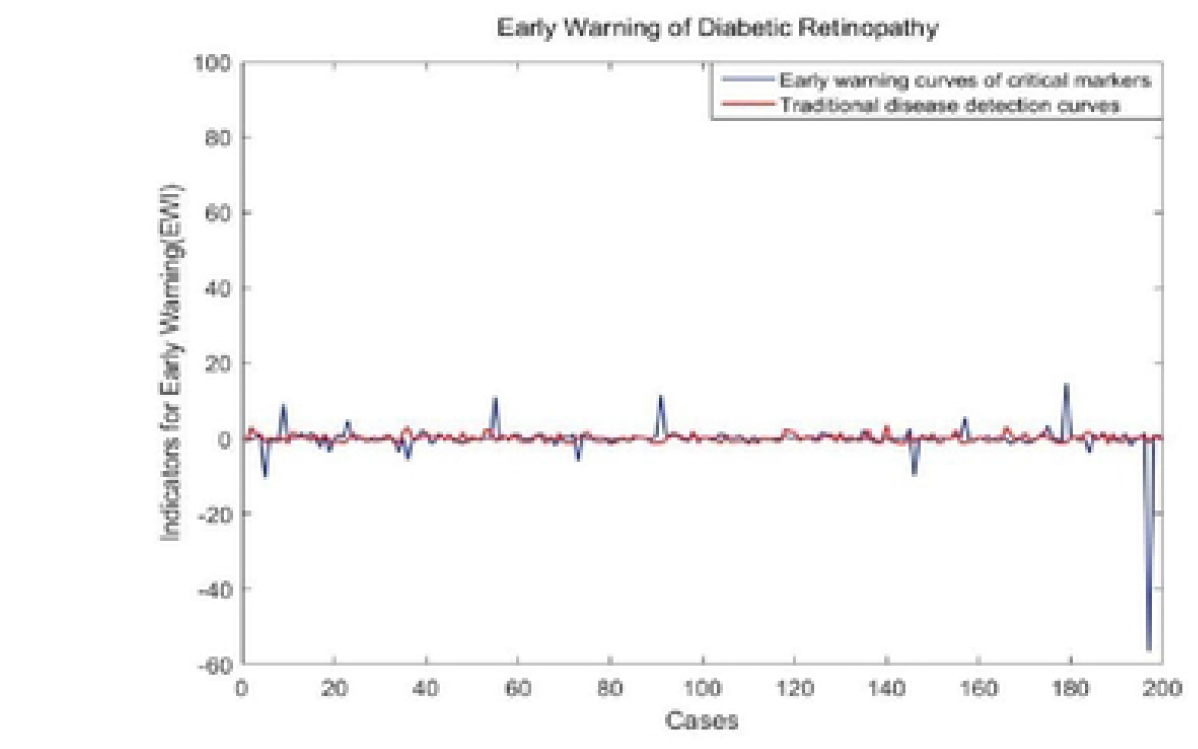
Correlation between key markers and non-key marker indicators

From **Figur**e 6-7, we can see that the correlation value of indicators in Figure 6 is higher on the whole, indicating that the key markers are more systematic and correlated, while spearman correlation coefficient in Figure 7 is lower on the whole with a small fluctuation range. Then we can conclude that the key markers are highly correlated and highly predictive. In the early warning of diabetic retinopathy, we can focus on the detection and research of key markers, which can save medical costs, and at the same time obtain a stronger warning than other indicators. This is a new and cheaper way to warn of diabetic retinopathy (complications).

### 3.4 Early warning of disease based on of key markers

Figure 8 shows the warning curve based on GLU data and the composite index of key markers respectively, in which red is the GLU fluctuation curve of 200 patients. If the traditional fasting blood glucose level is used to detect the occurrence of diabetes complications, the curve has a small trend of fluctuation, and it cannot play a good warning role for the occurrence of the disease. Abnormalities can only be detected after the onset of the disease. However, the composite index based on key markers to study disease early warning can show strong fluctuations during the individual’s medical testing phase, as shown in the blue curve. In this figure, there are six strong fluctuation signals, which indicate that the corresponding individual samples show abnormal physical signs, namely, the upcoming diabetic retinopathy based on this data set. Key markers are more sensitive than other indicators. And it doesn’t warn from a single perspective, it transmits warning signals to the occurrence of diseases from multiple aspects such as individual lipid metabolism, cell redox, influence on bone metabolism and bone resorption, and cell coagulation function.

**Figure 8.** Warning of diabetic retinopalhy based on key markers

## 4. Discussion

Diabetic retinopathy is one of the most common microvascular diseases in diabetes, which is essentially caused by abnormal blood glucose metabolism caused by insufficient insulin secretion or decreased activity. Its pathogenesis is complex, and it is affected by many factors in the development process. Therefore, in research, we should integrate multiple individual pathogenesis pathways. As a new way of disease warning, the key marker can detect the onset signal of the individual before the disease occurs. In this study, a novel algorithm was proposed based on BP neural network to detect the warning signal of diabetic retinopathy. This method not only saves the medical detecting cost, but also accelerates the medical efficiency of early warning and diagnosis.

In the paper, we applied the algorithm to the detection of key markers affecting pathogenesis and the early warning of disease onset. First, we introduced a disease warning method based on BP neural network, which studies how diabetic retinopathy is induced from the aspects of individual lipid metabolism, cell REDOX, bone metabolism and bone resorption, and cell clotting function. Second, according to individual medical test data, we identified key markers that induce diabetic retinopathy complications. In addition, the mechanism of action of the key markers was preliminarily determined, that is, the indicators in the key markers did not independently to induce the occurrence of the disease, but were highly correlated with each other. As shown in Figure 5, they coordinated and acted together before the occurrence of the disease. At the same time, when conducting disease early warning, key markers can serve as a complete module to form a strong early warning signal when a disease occurs. As shown in Figure 8, the key markers detected based on this algorithm have higher sensitivity and effectiveness in disease early warning. Finally, the disease warning method based on BP neural network provides a new method for the study of diabetic microvascular lesions. In clinical prediction and diagnosis, doctors can use key markers to give early warning of individual diseases and make early intervention.

The BP neural network used in this algorithm has a prominent ability to deal with complex relationships, and the network weight plays a very important role in the transmission process of nodes at each layer of the network. How to obtain, analyze and reasonably apply the weight matrix between each layer element is also a research feature of this paper. In this study, by adjusting the function and neuron parameters of BP neural network, the network model with the optimal training effect and the lowest experimental error was obtained. We can obtain the weight matrix under the network state, and perform hierarchical cluster analysis on the weight data of each medical index in the process of transmission. We select the molecular module with high weight and closest clustering distance as the key marker of disease early warning. We study both the independent importance of individual signs in disease early warning and the coordinated correlation between them in inducing disease occurrence. This method has unique advantages in the study of diabetes complications.

## 5. Conclusions

When studying disease early warning, traditional methods rely on linear model to study the linear relationship between two indexes. Based on the theory of BP neural network, in this paper, we studied the complex network relationship among the indexes that affect the occurrence of disease, and determined the key markers that induce the occurrence of diabetic retinopathy. In the study, we found that there was a strong correlation between key markers, while the correlation between internal indicators of key markers and non-marker indicators was low. It implies that indicators that become key markers in disease early warning are synergistic, which can form a complete module to transmit strong early warning signals before the occurrence of disease, and provide certain reference for clinical prediction and diagnosis. In studying the early warning of diabetic retinopathy, we found that clinical indicators that form key markers have been shown in many literatures to be closely related to the occurrence of the disease, such as glycosylated hemoglobin, serum uric acid, alanine aminotransferase, etc. They induce the occurrence of diseases from the aspects of individual lipid metabolism, cell redox, bone metabolism and bone resorption, as well as cell coagulation function. Therefore, this method can be used to detect key markers affecting diabetic complications, explore the induction mechanism of disease occurrence, and provide early warning for the occurrence of disease. Clinically, it is helpful for the intervention, diagnosis and treatment of diabetic complications. So. It can improve the quality of life of patients and reducing the mortality rate of diabetic complications.

## Data Availability

All relevant data are within the manuscript and its Supporting Information files.

## 7. Acknowledgments

The data came from the National Data Center for Population and Health Sciences. (https://www.ncmi.cn)

